# Construction and Validation of the Ohio Children’s Opportunity Index

**DOI:** 10.1101/2021.05.12.21257062

**Authors:** Naleef Fareed, Priti Singh, Pallavi Jonnalagadda, Christine M. Swoboda, Colin Odden, Nathan Doogan

**Affiliations:** CATALYST – The Center for the Advancement of Team Science, Analytics, and Systems Thinking, College of Medicine, The Ohio State University, Institute for Behavioral Medicine Research, 460 Medical Center Drive, Columbus, OH 43210, United States; Department of Biomedical Informatics, College of Medicine, The Ohio State University, Institute for Behavioral Medicine Research, 460 Medical Center Drive, Columbus, OH 43210, United States; Department of Family Medicine, College of Medicine, The Ohio State University, Institute for Behavioral Medicine Research, 460 Medical Center Drive, Columbus, OH 43210, United States; Department of Research Information Technology, College of Medicine, The Ohio State University, Suite 275, 530 W Spring St, Columbus, OH 43215, United States; Ohio Colleges of Medicine Government Resource Center, The Ohio State University, 1070 Carmack Road, Columbus, OH 43210, United States

**Keywords:** area-level measure, opportunity, children well-being, social determinants of health, neighborhood

## Abstract

**Objective:** To describe the development of an area-level measure of children’s opportunity, the Ohio Children’s Opportunity Index (OCOI).

**Data Sources/Study Setting:** Secondary data were collected from US census based-American Community Survey (ACS), US Environmental Protection Agency, US Housing and Urban Development, Ohio Vital Statistics, US Department of Agriculture-Economic Research Service, Ohio State University Center for Urban and Regional Analysis, Ohio Incident Based Reporting System, IPUMS National Historical Geographic Information System, and Ohio Department of Medicaid.

**Study Design:** OCOI domains were selected based on existing literature, which included family stability, infant health, children’s health, access, education, housing, environment and criminal justice domains. The composite index was developed using an equal weighting approach. Validation analyses were conducted between OCOI and health and race-related outcomes.

**Data Collection/Extraction Methods:** Data were aggregated at the census tract level.

**Principal Findings:** Composite OCOI scores ranged from 0-100 with an average value of 74.82 (SD, 17.00), was negatively skewed. Census tracts in the major metropolitan cities across Ohio represented 76% of the total census tracts in the least advantaged OCOI septile. OCOI served as a significant predictor of health and race-related outcomes. The average life expectancy at birth of children born in the most advantaged septile was approximately nine years more than those born in the least advantaged septile. Increases in OCOI were associated with decreases in proportion of Black (48 points lower in the most advantaged vs least advantaged septile), p<0.001) and Minority populations (54 points lower in most advantaged vs least advantaged septile, p<0.001).

**Conclusion:** As the first opportunity index developed for children in Ohio, the OCOI is a valuable resource for policy reform, especially related to health equity. Health care providers can use it to obtain more holistic views on their patients and implement interventions that can tackle barriers to childhood development.

## Introduction□

Unmet basic needs are likely to result in poor health outcomes across the lifespan^1,2^ making children living in poverty extremely vulnerable. Approximately 2.6 million of Ohio’s 11.5 million population are children. About 20% of these children in Ohio live in poverty, 16% are chronically absent from school, and 14-15% have a disability. Further, over 20,000 children in Ohio are homeless.^3^ Those who are most disadvantaged, while shouldering a disproportionately higher burden of poor health and risk factors for poor health, are also the least likely to access care when needed.^4^ Risk factors tend to cluster within individuals, families, and communities, worsening the inverse relationship between the need for healthcare and access to it.^5^ This phenomenon is apparent, for example, among vulnerable populations who have higher utilization of out-of-hours emergency health care rather than preventive health care, perpetuating the cycle of expensive, reactive care.^6^

The high infant mortality in Ohio,^7^ especially the wide disparity between infants born to White versus Black mothers□prompted the Ohio Department of Medicaid (ODM) to develop the Ohio Opportunity Index (OOI)^8^ and the Ohio Children’s Opportunity Index (OCOI) through the Infant Mortality Research Partnership and as a general movement to monitor deprivation among individuals from childhood and onwards. The objective was to aid the identification of deprived areas for targeted allocation of resources to improve health care delivery and health services, which has been shown to decrease disparities.^9^ Area-level indices of deprivation have been used in New Zealand and the United Kingdom not simply to study risk factors and outcomes but also for incorporation into healthcare delivery. □^10^

Individual factors only partially capture determinants of health and disease, drawing attention to the “place” effect^11–13^ - the social, economic, and physical conditions in the environment where people live, also called social determinants of health (SDoH).^14^ Several studies have formally decomposed the contributors of health outcomes into clinical care, health behaviors, socio-economic factors and physical environment.^15–18^ Characterizing the individual effect of any of these factors, particularly socioeconomic and environmental conditions that contribute between 20 and 50 percent to health outcomes, do not provide adequate guidance on how interventions or policies can be developed with greater precision for target populations.^17,19^ Hence, there is a need for nuance about modifiable attributes within a domain that can truly influence health outcomes.

The influence of SDoH vary based on the ecological level at which they operate. Poverty places a greater health burden on society than either of the leading behavioral risk factors —smoking or obesity.^20^ Individual poverty combined with living in an affluent neighborhood was not associated with negative health consequences, whereas living in a deprived neighborhood was associated with adverse health outcomes more so among poorer individuals, who may be more dependent on collective neighborhood resources.^21^ Deprivation is “a state of observable and demonstrable disadvantage relative to the local community or the wider society or nation,” and poverty on the other hand is the lack of resources to escape deprivation.^22^ An area-level deprivation index (ADI) reflects aggregate measures of SDoH at the neighborhood level.

Advances in computing power, geographic information systems (GIS), and statistical techniques like multi-level modeling allow for more sophisticated and detailed examination of area level SDoH than in the past.^23^ The Public Health Disparities Geocoding Project assessed a variety of single indicators and composite measures of socioeconomic deprivation and demonstrated gradients with outcomes like childhood lead poisoning, mortality, and low birth weight.^24,25^ Moreover, Krieger and colleagues demonstrated that indices of area level deprivation facilitated detection of larger socioeconomic gradients than more focused area level measures of education and wealth. Linking the area deprivation index with county-level mortality revealed widening inequalities in area level mortality on account of slower declines in mortality in deprived areas.^26^ These are but a few examples of the wealth of research suggesting that place matters.

Neighborhoods possess physical and social attributes that could affect health.^27^ Empirical research examining neighborhood effects on children and adolescents have established that there is considerable socioeconomic and racial segregation and that indicators like crime, social, and physical disorder tend to cluster at the neighborhood level.^28^ Predictors common to many childhood outcomes include concentrated poverty and racial isolation.^28^ Neighborhood disadvantage has been shown to be associated with child health outcomes such as behavioral problems and verbal ability.^29,30^ The influence of neighborhood can be recognized through the Moving to Opportunity Experiment. Moving to a more affluent neighborhood when children are younger than 13 was argued to have to an increase in college attendance and earnings.^31^ The seminal Whitehall studies have highlighted the social gradient or the socioeconomic differences in physical and mental illnesses and mortality.^9^

The OCOI is a measure of SDoH at the census tract level conveying opportunity information for children across the state of Ohio. We define children as anyone between birth and below the age of 18. As a neighborhood’s effect on children’s health is not exerted by a single factor but by a combination of them, the OCOI is a composite index of 53 neighborhood indicators spanning eight domains associated with healthy child development. The OCOI is not the first index associated with healthy childhood. The similarly-named Child Opportunity Index consists of 19 indicators corresponding to three domains: *educational, health and environment*, and *social and economic*. However, the Child Opportunity Index is only available for the U.S’ 100 largest metropolitan areas.^32^

In this article we describe the development of the OCOI. The purpose of the OCOI is to provide a measure of children’s opportunity in Ohio. Public health practitioners, policymakers, researchers, and healthcare providers can use the OCOI to identify neighborhoods of low and high opportunity in Ohio. In this article we first discuss the process of domain and input data selection followed by data extraction. Next, we discuss the four-step process involved in the construction of the OCOI based on seminal approaches.^33,34^ Finally, we report the association of the OCOI with life expectancy and proportions of minority populations to validate the index.

## Data and Methods

Measures of social determinants of children’s health and well-being were collected at the census tract level for Ohio. Census tracts are geographical sub-divisions of counties that contain an average of 4000 people.^3^ Because of similar neighborhood characteristics, federal and state agencies often collect tract level aggregates as a proxy for area-based information. Study data pertain to 2,940 tracts out of the total 2,952 census tracts in Ohio. Twelve tracts were excluded because of zero population.

Data were procured from federal sources such as the US census based-American Community Survey (ACS) data set, which is a freely available resource, and other state and federal agency administrative data sets (e.g., Medicaid claims and Department of Education school report card data). The Government Resource Center at the Ohio State University compiled the measures used for the construction of the OCOI. Information was gathered to represent the time period 2013-2017, inclusive.

### Domains and Variables

Deprivation indices are either represented by simple indicators measuring social deprivation alone, such as poverty,^35^ or as a composite score articulated from multiple mutually exclusive indicators or “domains”.^34^ Using the framework developed by Peter Townsend,^22^ the current study adopted a multi-dimensional and a multi-domain approach. The domains refer to a collection of constituent measures pertaining to economic, material, and psychosocial influences in humans. Additional details about these measures and associated attributes can be found in a study conducted by Pearce and colleagues.^36^ Guided by Townsend’s framework,^34^ the subject matter experts (maternal-child health and geospatial area deprivation measure development) and the study team identified a list of eight domains: *family stability, infant health, children’s health, access (to health care and food), education, housing, environment, and criminal justice* for OCOI construction. The study domains mostly overlap with the SDoH factors identified by the Center for Disease Control and Prevention (CDC) (life-enhancing resources such as food supply, housing, transportation, education, and health care) further substantiating their use.^37^ A brief description of the domains are as follows ^38,39^:

1. **Family Stability:** Measures early influences of family settings on children including family breakdown, parental relationship, and family income.
2. **Infant Health**: Determinants of children’s health that operate from before conception through birth. Maternal influences such as mother’s health, lifestyle, and social and physical environments have immediate effect on children’s health.
3. **Children’s Health:** Presence of chronic conditions in children that may affect their overall development.
4. **Access**: Poor geographical access to key local services
5. **Education**: Scholastic attainment and skills in local population that may lead to low health literacy.
6. **Housing**: Barriers to affordability of housing and stable housing conditions.
7. **Environment**: Physical space and characteristics, both natural and built, that influence health.
8. **Criminal Justice:** Likelihood for personal and material victimization at the local level.

The fifty-three constituent measures used in this study along with their corresponding data sources are listed in Table S1(Appendix A). These measures were summarized (within respective domains) to yield domain scores, which were further summarized to form the final OCOI. Figure 1 represents an outline of the process used to create the OCOI.

**Figure 1.**
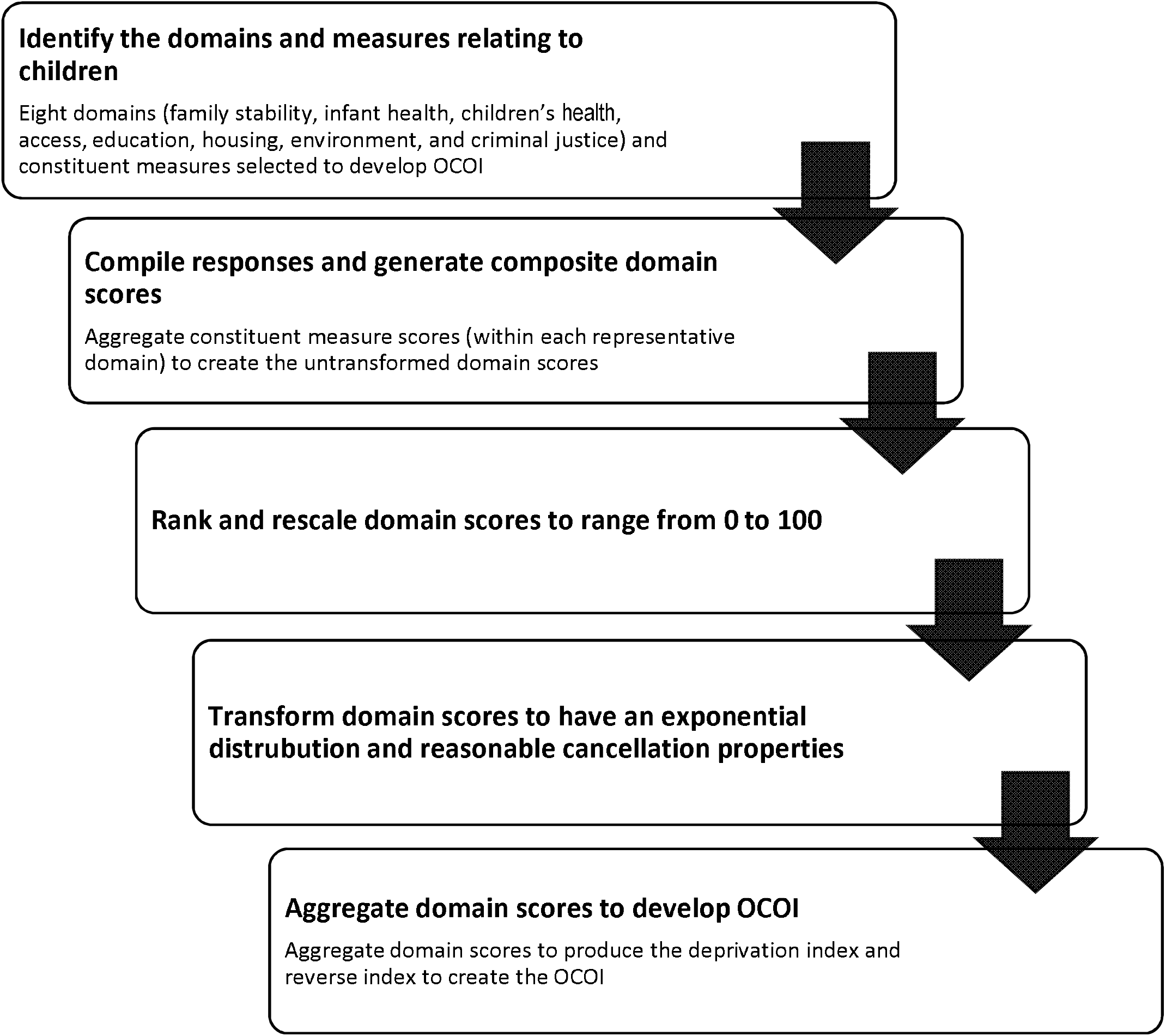
Flowchart representing steps involved in developing the Ohio Children’s Opportunity Index.

Of the 53 constituent measures, five were assigned to the family stability domain, seven to infant health, eight to children’s health, seven to access, eight to education, seven to housing, six to environment and five to the criminal justice domain. Some constituent measures were reverse coded to maintain a consistent direction with respect to what higher (opportunity) versus lower (opportunity) values mean.

### Validation Outcomes

The study-generated OCOI scores were tested for association with health-related outcomes previously linked to area-level deprivation.^40^ Probability of life expectancy was used as health outcome criterion for prediction based on OCOI. Life expectancy represents the expected average years of survival at birth. Data for this outcome was collected at the census tract level from 2010-2015 and retrieved from National Center for Health Statistics, CDC.^41^ Variability in population distribution, for Black and minority groups were also examined. Information regarding the percent of Blacks and minority population living within a tract were obtained from ACS.^42^

### Analysis

Statistical analyses were performed using R version 4.0.3. Raw data were obtained for measures across the 2,940 census tracts in Ohio. A multi-stage approach was adopted to generate the OCOI. First, univariate and bivariate analyses were conducted to explore the statistical distribution of variables, their missingness, and their relationship with other variables. Missing values were replaced with median values of the corresponding measure. Also, at this stage, we performed correlational analysis on the constituent measures to assess the grouping of the 53 variables (see supplementary Figure S1 for the correlation matrix). The next steps included a series of transformations to create a composite measure from raw scores. Following Townsend’s ^34^ and Noble’s^33^ approach, the OCOI was computed based on the following procedures:

1. Standardizing and averaging: Data were collected across 53 measures in different units such as proportions and counts. The first step in the analysis was to standardize these raw scores such that they have a mean of 0 and a standard deviation of 1 (i.e., *z*-scores). The standardized scores were averaged (within domains) to form domain scores and subsequently transformed in the following manner.
2. Ranking: The domain scores were ranked and scaled to range between zero and one (with the least deprived tract having a value 1/number of tracts).
3. Exponential distribution: The scaled rankings were then transformed to have an exponential distribution. According to Noble et al.,^33^ this helps each domain to have a common distribution, the same range, and identical maximum and minimum values of 0 and 100 respectively. The exponential distribution stretches out the distribution so that greater levels of deprivation score more highly. The transformed domain was given by Noble et al.,^33^ equation 1, wherein X is the transformed domain value, δ is a constant and R is the rank on the domain.

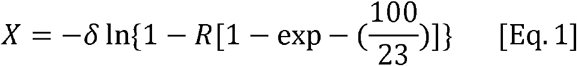
4. Equal weighting: These transformed final domain scores were then aggregated using a weighing technique. For this study we used the equal weighting method, wherein each domain was assigned a weight of 1/8 and aggregated to form the deprivation index. The equal weighting method is a seminal approach used by many European countries for calculating area-level deprivation scores.^43^ By doing so, we assume equal importance of all deprivation domains. This technique is known to produce valid area-level measures and significantly predict health outcomes such as mortality.^43^ The resultant tract-level scores represented deprivation index for Ohio and were reversed and scaled between 0 and 100 to create the OCOI for each census tract. Septiles were computed from the tract-level score to simplify interpretation.
5. Validation and sensitivity analysis: We used the same regression-based validation approach as previous studies to predict health-related outcomes.^43^ OCOI score categories (i.e., septiles) were used to predict life expectancy. The distribution of minority and Black population against OCOI categories were also examined using ordinary least squares (OLS) regression model specifications.

### OCOI Results

#### Univariate descriptive statistics

Table 1 presents descriptive statistics for the 53 constituent measures using aggregated data from 2013-2017. Out of 53 measures, responses for eleven measures were reversed to maintain a consistent direction: labor market engagement index, proportion of children with six or more well child primary care provider visits, proportion of children between ages three and six with one or more well-child primary care provider visits, low transportation cost index, behavioral health visits for children that meet access standards, proportion of primary care visits for children that meet the access standards of CMS, free lunch distribution, graduation rate, school performance index, schools value-added score, and environmental health hazard index.

**Table 1.**
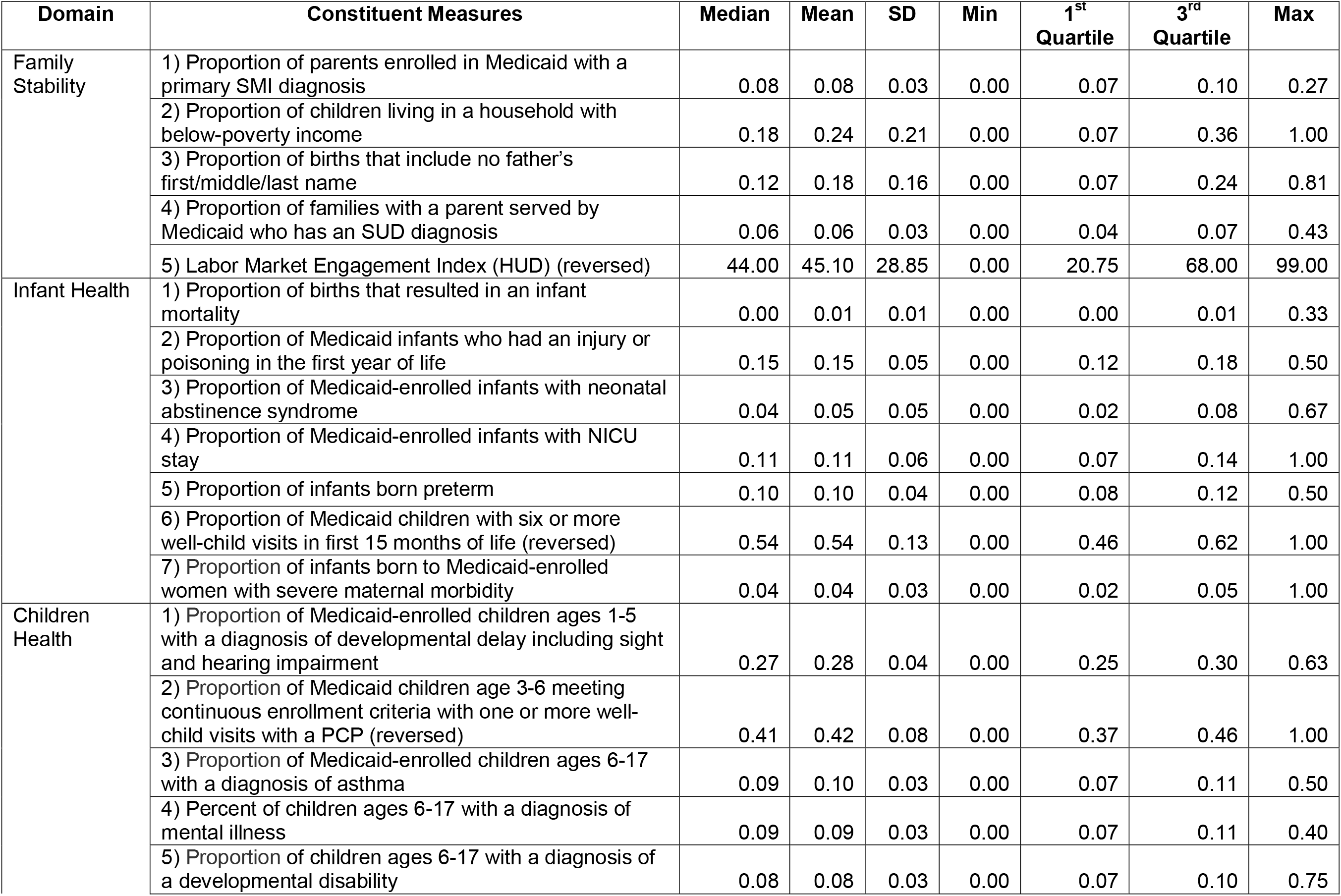
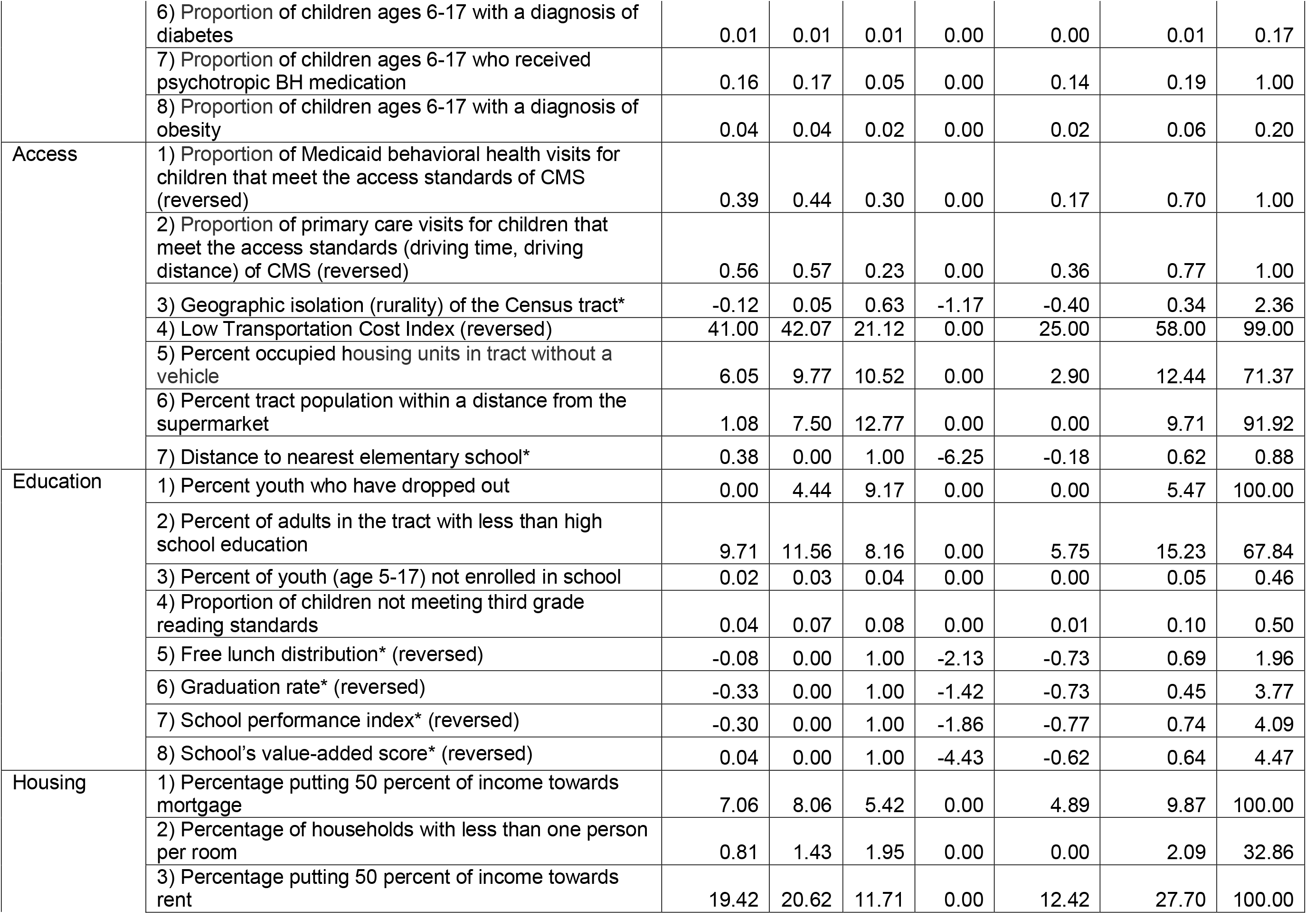
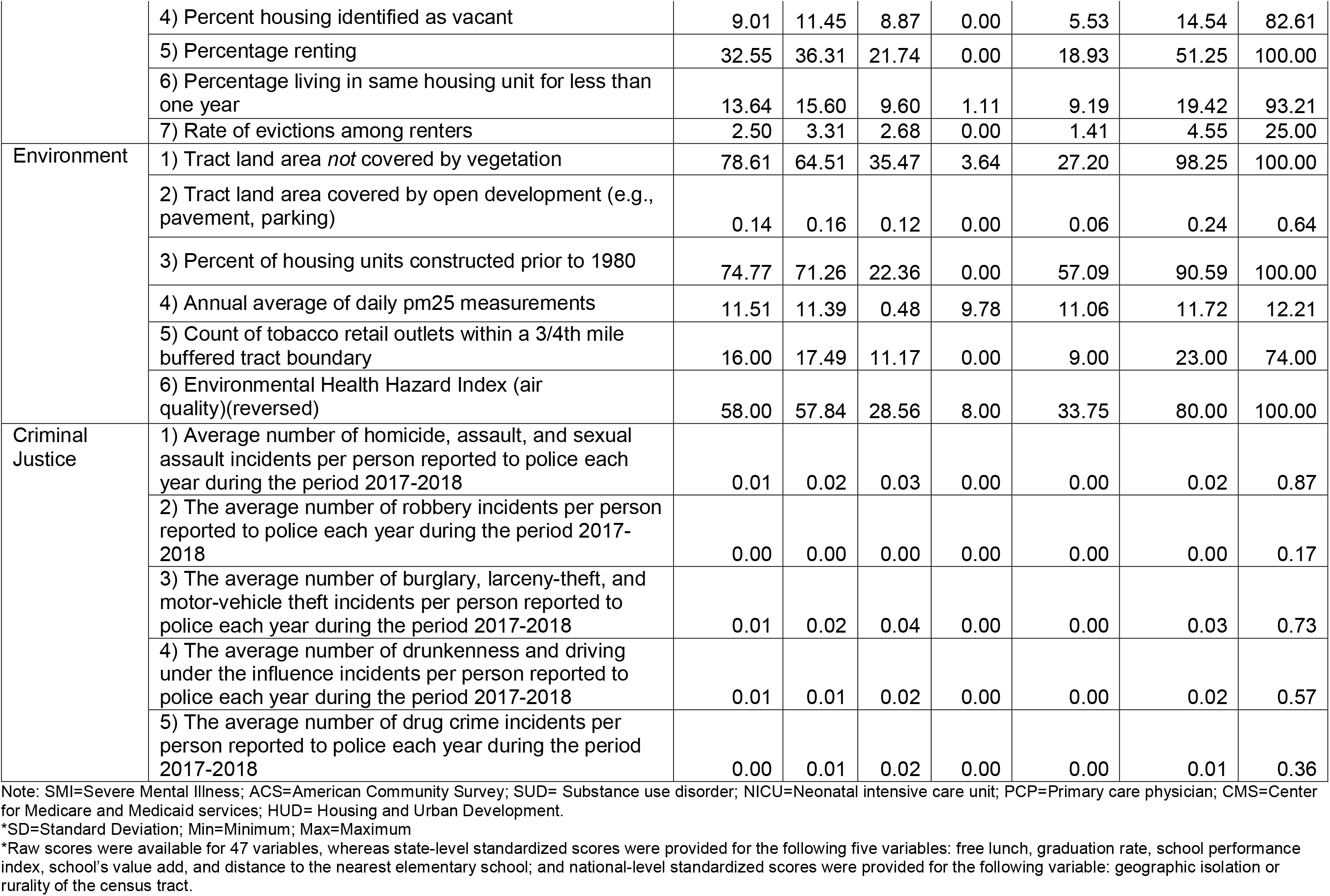
Univariate descriptive statistics of Ohio Children’s Opportunity Index variables.

| Domain | Constituent Measures | Median | Mean | SD | Min | 1<sup>st</sup> Quartile | 3<sup>rd</sup> Quartile | Max |
| --- | --- | --- | --- | --- | --- | --- | --- | --- |
| Family Stability | 1) Proportion of parents enrolled in Medicaid with a primary SMI diagnosis | 0.08 | 0.08 | 0.03 | 0.00 | 0.07 | 0.10 | 0.27 |
|  | 2) Proportion of children living in a household with below-poverty income | 0.18 | 0.24 | 0.21 | 0.00 | 0.07 | 0.36 | 1.00 |
|  | 3) Proportion of births that include no father's first/middle/last name | 0.12 | 0.18 | 0.16 | 0.00 | 0.07 | 0.24 | 0.81 |
|  | 4) Proportion of families with a parent served by Medicaid who has an SUD diagnosis | 0.06 | 0.06 | 0.03 | 0.00 | 0.04 | 0.07 | 0.43 |
|  | 5) Labor Market Engagement Index (HUD) (reversed) | 44.00 | 45.10 | 28.85 | 0.00 | 20.75 | 68.00 | 99.00 |
| Infant Health | 1) Proportion of births that resulted in an infant mortality | 0.00 | 0.01 | 0.01 | 0.00 | 0.00 | 0.01 | 0.33 |
|  | 2) Proportion of Medicaid infants who had an injury or poisoning in the first year of life | 0.15 | 0.15 | 0.05 | 0.00 | 0.12 | 0.18 | 0.50 |
|  | 3) Proportion of Medicaid-enrolled infants with neonatal abstinence syndrome | 0.04 | 0.05 | 0.05 | 0.00 | 0.02 | 0.08 | 0.67 |
|  | 4) Proportion of Medicaid-enrolled infants with NICU stay | 0.11 | 0.11 | 0.06 | 0.00 | 0.07 | 0.14 | 1.00 |
|  | 5) Proportion of infants born preterm | 0.10 | 0.10 | 0.04 | 0.00 | 0.08 | 0.12 | 0.50 |
|  | 6) Proportion of Medicaid children with six or more well-child visits in first 15 months of life (reversed) | 0.54 | 0.54 | 0.13 | 0.00 | 0.46 | 0.62 | 1.00 |
|  | 7) Proportion of infants born to Medicaid-enrolled women with severe maternal morbidity | 0.04 | 0.04 | 0.03 | 0.00 | 0.02 | 0.05 | 1.00 |
| Children Health | 1) Proportion of Medicaid-enrolled children ages 1-5 with a diagnosis of developmental delay including sight and hearing impairment | 0.27 | 0.28 | 0.04 | 0.00 | 0.25 | 0.30 | 0.63 |
|  | 2) Proportion of Medicaid children age 3-6 meeting continuous enrollment criteria with one or more well-child visits with a PCP (reversed) | 0.41 | 0.42 | 0.08 | 0.00 | 0.37 | 0.46 | 1.00 |
|  | 3) Proportion of Medicaid-enrolled children ages 6-17 with a diagnosis of asthma | 0.09 | 0.10 | 0.03 | 0.00 | 0.07 | 0.11 | 0.50 |
|  | 4) Percent of children ages 6-17 with a diagnosis of mental illness | 0.09 | 0.09 | 0.03 | 0.00 | 0.07 | 0.11 | 0.40 |
|  | 5) Proportion of children ages 6-17 with a diagnosis of a developmental disability | 0.08 | 0.08 | 0.03 | 0.00 | 0.07 | 0.10 | 0.75 |
|  | 6) Proportion of children ages 6-17 with a diagnosis of diabetes | 0.01 | 0.01 | 0.01 | 0.00 | 0.00 | 0.01 | 0.17 |
|  | 7) Proportion of children ages 6-17 who received psychotropic BH medication | 0.16 | 0.17 | 0.05 | 0.00 | 0.14 | 0.19 | 1.00 |
|  | 8) Proportion of children ages 6-17 with a diagnosis of obesity | 0.04 | 0.04 | 0.02 | 0.00 | 0.02 | 0.06 | 0.20 |
| Access | 1) Proportion of Medicaid behavioral health visits for children that meet the access standards of CMS (reversed) | 0.39 | 0.44 | 0.30 | 0.00 | 0.17 | 0.70 | 1.00 |
|  | 2) Proportion of primary care visits for children that meet the access standards (driving time, driving distance) of CMS (reversed) | 0.56 | 0.57 | 0.23 | 0.00 | 0.36 | 0.77 | 1.00 |
|  | 3) Geographic isolation (rurality) of the Census tract* | -0.12 | 0.05 | 0.63 | -1.17 | -0.40 | 0.34 | 2.36 |
|  | 4) Low Transportation Cost Index (reversed) | 41.00 | 42.07 | 21.12 | 0.00 | 25.00 | 58.00 | 99.00 |
|  | 5) Percent occupied housing units in tract without a vehicle | 6.05 | 9.77 | 10.52 | 0.00 | 2.90 | 12.44 | 71.37 |
|  | 6) Percent tract population within a distance from the supermarket | 1.08 | 7.50 | 12.77 | 0.00 | 0.00 | 9.71 | 91.92 |
|  | 7) Distance to nearest elementary school* | 0.38 | 0.00 | 1.00 | -6.25 | -0.18 | 0.62 | 0.88 |
| Education | 1) Percent youth who have dropped out | 0.00 | 4.44 | 9.17 | 0.00 | 0.00 | 5.47 | 100.00 |
|  | 2) Percent of adults in the tract with less than high school education | 9.71 | 11.56 | 8.16 | 0.00 | 5.75 | 15.23 | 67.84 |
|  | 3) Percent of youth (age 5-17) not enrolled in school | 0.02 | 0.03 | 0.04 | 0.00 | 0.00 | 0.05 | 0.46 |
|  | 4) Proportion of children not meeting third grade reading standards | 0.04 | 0.07 | 0.08 | 0.00 | 0.01 | 0.10 | 0.50 |
|  | 5) Free lunch distribution* (reversed) | -0.08 | 0.00 | 1.00 | -2.13 | -0.73 | 0.69 | 1.96 |
|  | 6) Graduation rate* (reversed) | -0.33 | 0.00 | 1.00 | -1.42 | -0.73 | 0.45 | 3.77 |
|  | 7) School performance index* (reversed) | -0.30 | 0.00 | 1.00 | -1.86 | -0.77 | 0.74 | 4.09 |
|  | 8) School's value-added score* (reversed) | 0.04 | 0.00 | 1.00 | -4.43 | -0.62 | 0.64 | 4.47 |
| Housing | 1) Percentage putting 50 percent of income towards mortgage | 7.06 | 8.06 | 5.42 | 0.00 | 4.89 | 9.87 | 100.00 |
|  | 2) Percentage of households with less than one person per room | 0.81 | 1.43 | 1.95 | 0.00 | 0.00 | 2.09 | 32.86 |
|  | 3) Percentage putting 50 percent of income towards rent | 19.42 | 20.62 | 11.71 | 0.00 | 12.42 | 27.70 | 100.00 |
|  | 4) Percent housing identified as vacant | 9.01 | 11.45 | 8.87 | 0.00 | 5.53 | 14.54 | 82.61 |
|  | 5) Percentage renting | 32.55 | 36.31 | 21.74 | 0.00 | 18.93 | 51.25 | 100.00 |
|  | 6) Percentage living in same housing unit for less than one year | 13.64 | 15.60 | 9.60 | 1.11 | 9.19 | 19.42 | 93.21 |
|  | 7) Rate of evictions among renters | 2.50 | 3.31 | 2.68 | 0.00 | 1.41 | 4.55 | 25.00 |
| Environment | 1) Tract land area *not* covered by vegetation | 78.61 | 64.51 | 35.47 | 3.64 | 27.20 | 98.25 | 100.00 |
|  | 2) Tract land area covered by open development (e.g., pavement, parking) | 0.14 | 0.16 | 0.12 | 0.00 | 0.06 | 0.24 | 0.64 |
|  | 3) Percent of housing units constructed prior to 1980 | 74.77 | 71.26 | 22.36 | 0.00 | 57.09 | 90.59 | 100.00 |
|  | 4) Annual average of daily pm25 measurements | 11.51 | 11.39 | 0.48 | 9.78 | 11.06 | 11.72 | 12.21 |
|  | 5) Count of tobacco retail outlets within a 3/4th mile buffered tract boundary | 16.00 | 17.49 | 11.17 | 0.00 | 9.00 | 23.00 | 74.00 |
|  | 6) Environmental Health Hazard Index (air quality)(reversed) | 58.00 | 57.84 | 28.56 | 8.00 | 33.75 | 80.00 | 100.00 |
| Criminal Justice | 1) Average number of homicide, assault, and sexual assault incidents per person reported to police each year during the period 2017-2018 | 0.01 | 0.02 | 0.03 | 0.00 | 0.00 | 0.02 | 0.87 |
|  | 2) The average number of robbery incidents per person reported to police each year during the period 2017-2018 | 0.00 | 0.00 | 0.00 | 0.00 | 0.00 | 0.00 | 0.17 |
|  | 3) The average number of burglary, larceny-theft, and motor-vehicle theft incidents per person reported to police each year during the period 2017-2018 | 0.01 | 0.02 | 0.04 | 0.00 | 0.00 | 0.03 | 0.73 |
|  | 4) The average number of drunkenness and driving under the influence incidents per person reported to police each year during the period 2017-2018 | 0.01 | 0.01 | 0.02 | 0.00 | 0.00 | 0.02 | 0.57 |
|  | 5) The average number of drug crime incidents per person reported to police each year during the period 2017-2018 | 0.00 | 0.01 | 0.02 | 0.00 | 0.00 | 0.01 | 0.36 |

Note: SMI=Severe Mental Illness; ACS=American Community Survey; SUD= Substance use disorder; NICU=Neonatal intensive care unit; PCP=Primary care physician; CMS=Center for Medicare and Medicaid services; HUD= Housing and Urban Development.
\*SD=Standard Deviation; Min=Minimum; Max=Maximum
\*Raw scores were available for 47 variables, whereas state-level standardized scores were provided for the following five variables: free lunch, graduation rate, school performance index, school's value add, and distance to the nearest elementary school; and national-level standardized scores were provided for the following variable: geographic isolation or rurality of the census tract.

### OCOI scores

Graphical distribution and descriptive statistics for the study-generated OCOI scores are reported in Figure S2 and Table 2 respectively. As shown by the histogram, the distribution of OCOI scores displayed a negative skew, indicating higher opportunities for children in Ohio for most tracts compared to normally distributed outcomes. The average OCOI score was 74.82 (SD, 17.00).

**Table 2.** Summary of Ohio Children’s Opportunity Index scores and domains.

|  | **Median** | **Mean** | **SD** | **Min** | **1st**<br>**Quartile** | **3rd**<br>**Quartile** | **Max** |
| --- | --- | --- | --- | --- | --- | --- | --- |
| OCOI | 79.35 | 74.82 | 17.00 | 0.00 | 65.64 | 87.84 | 100 |
| Septile 1 | 44.01 | 42.49 | 9.50 | 0.00 | 37.10 | 49.99 | 54.30 |
| Septile 2 | 62.59 | 62.01 | 4.20 | 54.31 | 58.27 | 65.62 | 68.41 |
| Septile 3 | 72.47 | 72.50 | 2.31 | 68.42 | 70.47 | 74.48 | 76.32 |
| Septile 4 | 79.35 | 79.23 | 1.64 | 76.32 | 77.78 | 80.50 | 82.12 |
| Septile 5 | 84.50 | 84.53 | 1.37 | 82.12 | 83.40 | 84.57 | 86.73 |
| Septile 6 | 88.85 | 88.90 | 1.26 | 86.73 | 87.83 | 89.92 | 91.09 |
| Septile 7 | 93.68 | 94.10 | 2.10 | 91.1 | 92.34 | 95.56 | 100 |

Figure 2 presents a choropleth map of the OCOI across the state census tracts. Census tracts in the metropolitan cities of Cleveland, Columbus, Cincinnati, Toledo, and Dayton contained 29.28%, 15.47%, 13.33%, 9.4%, and 8.09% (together a total of 75.7%) of the total census tracts in the lowest OCOI septile (Q1), respectively.

**Figure 2.**
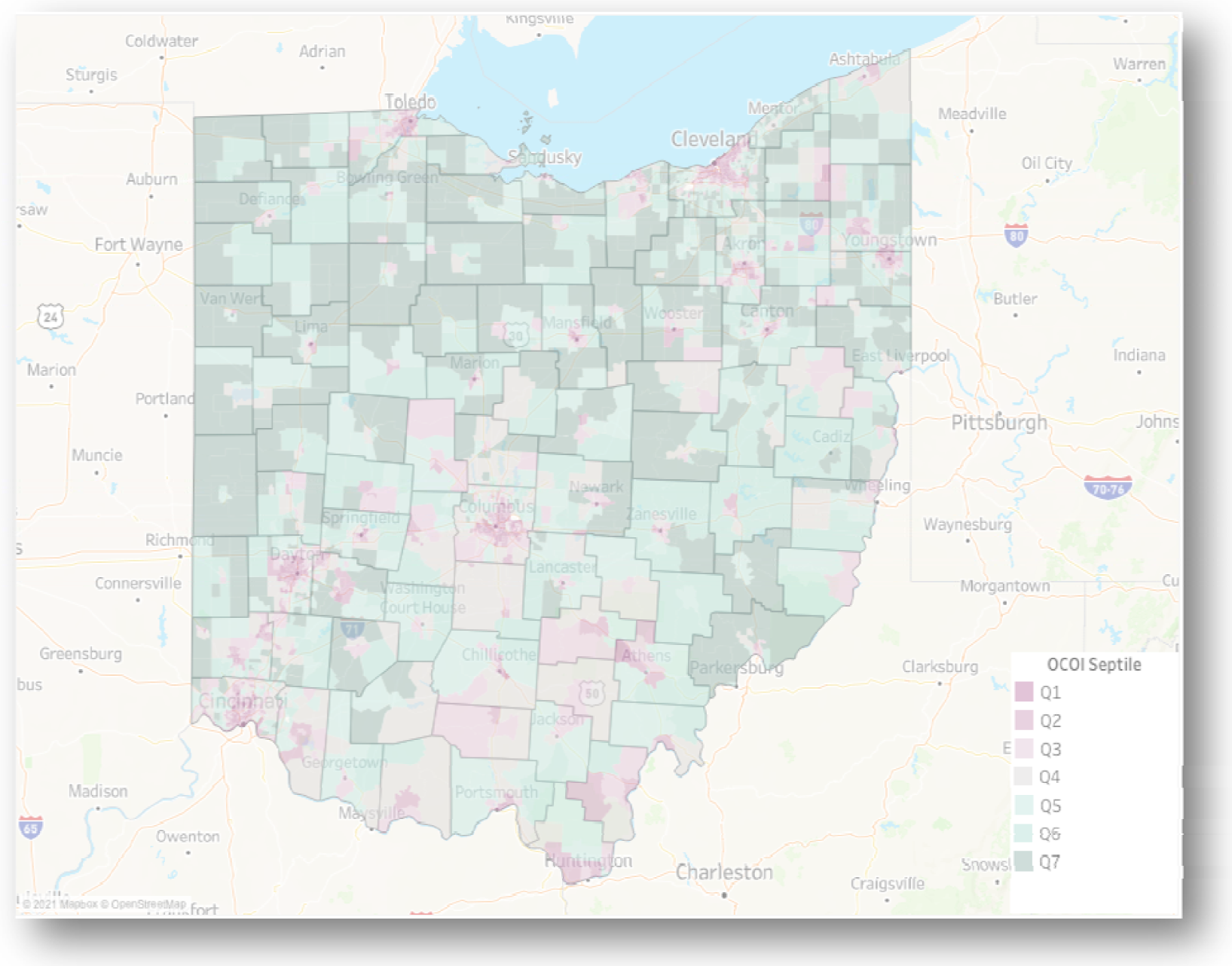
Ohio Children’s Opportunity Index scores on a choropleth map of Ohio. Ohio Children’s Opportunity Index (OCOI) distribution (as septiles of scores) displayed across tracts and counties Q1 represents least advantaged census tracts.

Figure 3 Illustrates patterns in OCOI scores within a single neighborhood in Columbus, Ohio. Upper Arlington anecdotally represents a neighborhood of high opportunity and living standards. There are 46% of tracts in Upper Arlington in the top three OCOI septiles, albeit 26% of the tracts in this neighborhood are in the bottom three septiles. The tracts with low OCOI scores in this neighborhood reflect a strong contrast to the immediately adjacent tracts in regard to domains such as children’s health and family stability.

**Figure 3.**
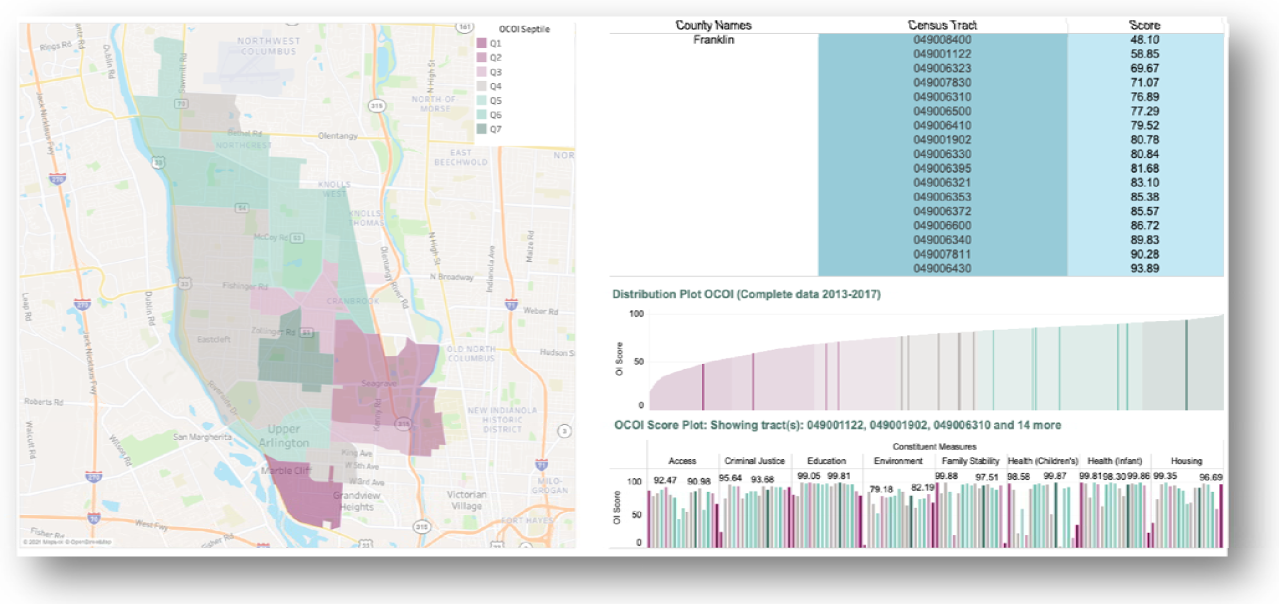
Illustrating distribution of Ohio Children’s Opportunity Index scores using a neighborhood view. Ohio Children’s Opportunity Index (OCOI) distribution (as septiles of scores) and sub-domain scores displayed across tracts for one neighborhood in Columbus, Ohio. Q1 represents least advantaged census tracts.

### Validation Results

We validated our OCOI measure using regression analysis wherein OCOI scores (collapsed to septiles, with the lowest used as the reference) were used as predictors for reference measures: health-related outcomes and neighborhood proportion of minority populations. OCOI scores were a significant predictor of each reference measure (p<0.001). The average life expectancy of children born in septile 7 was approximately 9 years more than those born in septile 1. The variability in Black and Minority population on OCOI scores was also examined based on OCOI septiles. The percentage of Black population living in septile 7 was 48 points lesser than those in septile 1. Likewise, the percentage of Minority population living in septile 7 was 54 points lesser than septile 1. From our test of trends, we found that trends were present in health outcomes and population distribution across the ordered levels or septiles of OCOI (p<0.001). See supplementary Table S2 for estimates from the validation analyses.

## Discussion

The OCOI was created to codify the geographic distribution of SDoH in the state of Ohio, particularly those that are likely to impact infants and children. The final OCOI was made up of eight domains comprising 53 variables that vary geographically. Analysis of the OCOI by census tract also shows that it captures variations by census tract that may be missed at coarser geographies. We found that increases in the OCOI scores were associated with higher tract-average life expectancy and minority population proportion.

The OCOI was inspired by the Ohio Opportunity Index (OOI), a related index that describes general deprivation of geographic areas in Ohio and consists of a different set of variables and domains. The researchers and stakeholders who developed the OOI realized that there were factors affecting children’s health and development that do not affect adults in the same way, along with factors in the OOI that do not influence children as much, motivating the development of a more specific index for children. The OCOI domains of family stability, environment, infant health, and children’s health are not in the OOI, but are important predictors of children’s health because they are associated with adolescent and adult health, social, and educational development.^44–48^ There is significant evidence that children living in more deprived areas are more likely to experience poor social, behavioral, health, and economic outcomes not only in childhood but throughout life, highlighting the importance of a children’s index.^49–52^ Moreover, both indices are based on data that contains information specific to Ohio and its population.

There is conflicting evidence regarding the extent to which children’s outcomes are affected by poverty on the family level versus the neighborhood level. Some research shows children in poor families may experience worse outcomes, a form of “double disadvantage,” when they live and attend school alongside more affluent versus similarly positioned peers as opposed to those who live near peers in similar levels of poverty.^53^ Other research, including data from the Moving to Opportunity (MTO) Study, provides conflicting evidence that growing up in better quality neighborhoods can improve the adult earnings of low-income children that move out of more deprived areas.^31^ The OCOI shows that many of the census tracts with the lowest opportunity are in urban areas, where children may be very close to tracts with extremely different OCOI scores, which could negatively affect their subjective social status.^53^ In addition, there are some possible negative effects of moving children from a low opportunity area to a high opportunity area other than their comparatively low social status, including low academic achievement ^54^ and antisocial behavior.^55^ Efforts to improve OCOI scores should focus on providing resources and helping areas with the most deprivation to increase equality of opportunity, rather than moving children out of low opportunity areas at the expense of the children who remain in them.

This is the first children’s opportunity index developed for the state of Ohio, however, there are similar efforts to map children’s deprivation or opportunity in other parts of the United States. There is a national Children’s Opportunity Index that uses data for the 100 most populated metro areas in the United States, however, individual domains are not shown and only metropolitan areas are shown.^56^ This index has been widely used for metropolitan areas, but many rural, suburban, and areas near small cities were excluded. The Opportunity Atlas is a national index that shows the likelihood of a child in different census tracts experiencing certain economic and educational outcomes as an adult. This contrasts with the OCOI because it focuses more on economic outcomes including income, employment, graduation rates, and other individual variables rather than health. Additionally, it focuses on the likelihood of adult outcomes but not problems that may affect the children living in those areas in the present, such as crime or family stability, which could influence stress and other mediators of those later outcomes. Other countries or groups of countries including South Africa ^57^ and the European Union ^58^ have also created deprivation or opportunity indices for children, and some states in the United States have limited indices studying childhood poverty alone,^59,60^ but no sources were found for multidimensional child deprivation indices on a state or regional level. Studying child deprivation on a smaller scale offers state and regional governments the ability to allocate funding for specific interventions on a local level.

The OCOI showed a significant relationship between percentage of minorities in a census tract and overall scores, with higher minority populations associated with lower scores. This is similar to results from the National Opportunity Index, with their index showing across 100 metro regions Child Opportunity Scores for White children (score=73) that were higher than for Black (score=24) or Hispanic (score=33) children.^61^ There is a complex relationship between race, geography, and deprivation in the United States due to segregation, discrimination, White flight, redlining, and institutional racism.^62^ The poverty rates for Black and Hispanic children are more than double that of White children in the United States.^63,64^ The compounding of low family wealth and living in deprived regions make it even less likely for minority children to escape poverty. It was found that upward mobility, defined by a child in the lowest income quintile reaching the highest as an adult, was greatest for areas with less segregation, less income inequality, better schools, greater social capital, and more family stability.^65^ Investing in the most deprived areas by improving education and decreasing income inequality may help alleviate some of the effects that are continuing to disproportionately hurt minority neighborhoods.^49^

The OCOI has potential for improvements that may further enhance its ability to communicate deprivation information. The researchers are working on visual tools and dashboards that describe and map OCOI and the individual domains. These visual tools could potentially incorporate race and ethnicity information to show the compounding of race and deprivation. These tools will allow researchers, public health, and government initiatives map areas to target for interventions, and learn more specifically which resources may be most needed in low opportunity areas. Additional years of data are being added as they become available, and will assist researchers in seeing changes in trends over time. Linking this change data with outcome data will enable researchers to study whether public programs and initiatives affected change in deprived areas and further inform decisions regarding specific resources needed.

## Limitations

The OCOI has some limitations that affect its scope and intended use. There is individual variation in deprivation within census tracts, and these measures should not be used alone to infer an individual’s risks. The use of varied data sources, especially census data, which is updated on 10-year cycles, may limit the frequency of data updates, and some variables may update at different rates than others. The reference time period of 2013-2017 glosses how conditions change over time, and the OCOI is not designed to capture the role of neighborhood change in health.^66^ Those using deprivation measures should remain cautious when interpreting what “low opportunity” means and make efforts to not promote negative characterizations of neighborhoods that need help. The intention is to help reduce inequity. However, if resources are allocated improperly, tools like the OCOI could even further divide communities. For example, if the OCOI is used by businesses or housing developers to find higher opportunity areas and avoid lower opportunity areas, they may continue investing in and improving places that do not need help. Policymakers should use these tools to specifically target areas and outcomes that need the most help, and also be careful to not waste resources on interventions that are unnecessary based on the data.

## Conclusion

Health is multifaceted and influenced by a constellation of physical, environmental, social, and economic factors that in turn interact with individual characteristics.

Generally, individuals residing in more deprived areas suffer worse health outcomes and measuring area-level conditions is an important contribution to identifying and addressing health disparities. Collaboration between health and social services in these lower opportunity areas should be encouraged to address multiple needs. Children born and raised in more deprived neighborhoods may have more health, social, behavioral, and economic problems as adolescents and adults,^49–52^ and are less likely to escape their low economic positions than children with similar family socioeconomic status living in more affluent areas.^66^ The factors affecting the health of adults and children differ somewhat, highlighting the importance of deprivation measures targeted specifically for younger populations. Area-level measures like the OCOI can help public health efforts more effectively map where the highest need is and which specific interventions would be beneficial.

## Protection of Human and Animal Subjects

Our study was reviewed by our IRB and deemed exempt.

## Supporting information

see supplementary

## Data Availability

Data used for this manuscript consists of publicly available data and restricted data.

https://childrensopportunityindex.github.io/The-Ohio-Opportunity-Index-Project/

