## Supplementary material for "Construction and Validation of the Ohio Children’s Opportunity Index": see supplementary

**Supplementary Table**

**Table S1. Ohio Children’s Opportunity Index domains, constituent measures, and data sources**

| **Domain** | **Constituent Measures** | **Data Source** |
| --- | --- | --- |
| Family Stability | 1) Proportion of parents enrolled in Medicaid with a primary SMI diagnosis | Medicaid Admin |
|  | 2) Proportion of children living in a household with below-poverty income | ACS |
|  | 3) Proportion of births that include no father’s first/middle/last name | VS births |
|  | 4) Proportion of families with a parent served by Medicaid who has an SUD diagnosis | Medicaid Admin |
|  | 5) Labor Market Engagement Index (HUD)(reversed) | HUD |
| Infant Health | 1) Proportion of births that resulted in an infant mortality | VS Births, Deaths |
|  | 2) Proportion of Medicaid infants who had an injury or poisoning in the first year of life | Medicaid Admin |
|  | 3) Medicaid-enrolled infants with neonatal abstinence syndrome | Medicaid Admin |
|  | 4) Medicaid-enrolled infants with NICU stay | Medicaid Admin |
|  | 5) Proportion of infants born preterm | VS Births |
|  | 6) Medicaid children with six or more well-child visits in first 15 months of life (reversed) | Medicaid Admin |
|  | 7) Proportion of infants born to Medicaid-enrolled women with severe maternal morbidity | Medicaid Admin |
| Children’s Health (non-infant) | 1) Proportion of Medicaid-enrolled children ages 1-5 with a diagnosis of developmental delay including sight and hearing impairment | Medicaid Admin |
|  | 2) Proportion of Medicaid children age 3-6 meeting continuous enrollment criteria with one or more well-child visits with a PCP (reversed) | Medicaid Admin |
|  | 3) Proportion of Medicaid-enrolled children ages 6-17 with a diagnosis of asthma | Medicaid Admin |
|  | 4) Proportion of Medicaid-enrolled children ages 6-17 with a diagnosis of mental illness | Medicaid Admin |
|  | 5) Proportion of Medicaid enrolled children ages 6-17 with a diagnosis of a developmental disability | Medicaid Admin |
|  | 6) Proportion of Medicaid enrolled children ages 6-17 with a diagnosis of diabetes | Medicaid Admin |
|  | 7) Proportion of Medicaid enrolled children ages 6-17 who received psychotropic BH medication | Medicaid Admin |
|  | 8) Proportion of Medicaid-enrolled children ages 6-17 with a diagnosis of obesity | Medicaid Admin |
| Access | 1) Proportion of Medicaid behavioral health visits for children that meet the access standards of CMS (reversed) | Medicaid Admin |
|  | 2) Proportion of primary care visits for children that meet the access standards (driving time, driving distance) of CMS (reversed) | Medicaid Admin |
|  | 3) Geographic isolation (rurality) of the Census tract | Doogan et al.,^67^ 2018 |
|  | 4) Low Transportation Cost Index | HUD |
|  | 5) Percent occupied housing units in tract without a vehicle | ACS |
|  | 6) Percent tract population within a distance from the supermarket | USDA-ERS |
|  | 7) Distance to nearest elementary school | CURA |
| Education | 1) Percent youth who have dropped out | ACS |
|  | 2) Percent of adults in the tract with more than high school education | ACS |
|  | 3) Percent of youth (age 5-17) not enrolled in school | ACS |
|  | 4) Proportion of children not meeting third grade reading standards | ACS |
|  | 5) Free lunch distribution (reversed) | ACS |
|  | 6) Graduation rate (reversed) | ACS |
|  | 7) School performance index (reversed) | ODE |
|  | 8) School’s value-added score (reversed) | ODE |
| Housing | 1) Percentage putting 50 percent of income towards mortgage | ACS |
|  | 2) Percentage of households with less than one person per room | ACS |
|  | 3) Percentage putting 50 percent of income towards rent | ACS |
|  | 4) Percent housing identified as vacant | ACS |
|  | 5) Percent renting | ACS |
|  | 6) Percentage living in same housing unit for less than one year | ACS |
|  | 7) Rate of evictions among renters | Eviction lab |
| Environment | 1) Tract land area *not* covered by vegetation | NHGIS |
|  | 2) Tract land area covered by open development (e.g., pavement, parking) | NHGIS |
|  | 3) Percent of housing units constructed prior to 1980 | ACS |
|  | 4) Annual average of daily pm25 measurements | EPA |
|  | 5) Count of tobacco retail outlets within a 3/4^th^ mile buffered tract boundary | Burgoon et al.,^68^ 2019 |
|  | 6) Environmental Health Hazard Index (air quality) | HUD |
| Criminal Justice | 1) Average number of homicide, assault, and sexual assault incidents per person reported to police each year during the period 2017-2018 | OIBRS |
|  | 2) The average number of robbery incidents per person reported to police each year during the period 2017-2018 | OIBRS |
|  | 3) The average number of burglary, larceny-theft, and motor-vehicle theft incidents per person reported to police each year during the period 2017-2018 | OIBRS |
|  | 4) The average number of drunkenness and driving under the influence incidents per person reported to police each year during the period 2017-2018 | OIBRS |
|  | 5) The average number of drug crime incidents per person reported to police each year during the period 2017-2018 | OIBRS |

Note: SMI=Severe Mental Illness; ACS=American Community Survey; VS=Vital statistics; SUD= Substance use disorder; HUD= Housing and Urban Development; NICU=Neonatal intensive care unit; PCP=Primary care physician; BH=Behavioral Health; USDA_ERS=U.S. Department of Agriculture-Economic Research Service; CMS=Centers for Medicare and Medicaid services; CURA=Center for Urban and Regional Analysis Ohio State University; ODE=Ohio Department of Education; EPA=Environmental Protection Agency; NHIGS=National Historical Geographic Information System; OIBRS=Ohio Incident Based Reporting System.

**Table S2. Unadjusted results of regression of key outcome and racial variables and Ohio Children’s Opportunity Index (OCOI) septiles**

| **OCOI septiles**  (Septile 1=reference) | **Life expectancy**  **(Adj. R^2 =^0.49)** | **Black Population** (Proportion)  **(Adj. R^2 =^0.49)** | **Minority population** (Proportion)  **(Adj. R^2 =^0.47)** |
| --- | --- | --- | --- |
| Septile 2 | 2.41(0.21)*** | -0.19(0.01)*** | -0.20(0.01)*** |
| Septile 3 | 4.69(0.21)*** | -0.35(0.01)*** | -0.38(0.01)*** |
| Septile 4 | 6.51(0.21)*** | -0.42(0.01)*** | -0.46(0.01)*** |
| Septile 5 | 6.80(0.21)*** | -0.45(0.01)*** | -0.50(0.01)*** |
| Septile 6 | 7.94(0.21)*** | -0.47(0.01)*** | -0.52(0.01)*** |
| Septile 7 | 8.78(0.20)*** | -0.48(0.01)*** | -0.54(0.01)*** |

Regression Coefficient (Standard Error), ***p<0.001, Adj. R^2^=Adjusted R-squared.

Q1 represents least advantaged census tracts.

**Supplementary Figures**

**Figure S1**

**Correlation matrix heatmap Ohio Children’s Opportunity Index constituent measures**

Figure S1. represents the bi-variate correlation between the constituent measures as a color-coded matrix. Measures with similar outcomes are represented by red color (denser the color, stronger the relationship) while unrelated (no relationship) or dissimilar (opposite relationship) domains are represented by white and blue colors, respectively

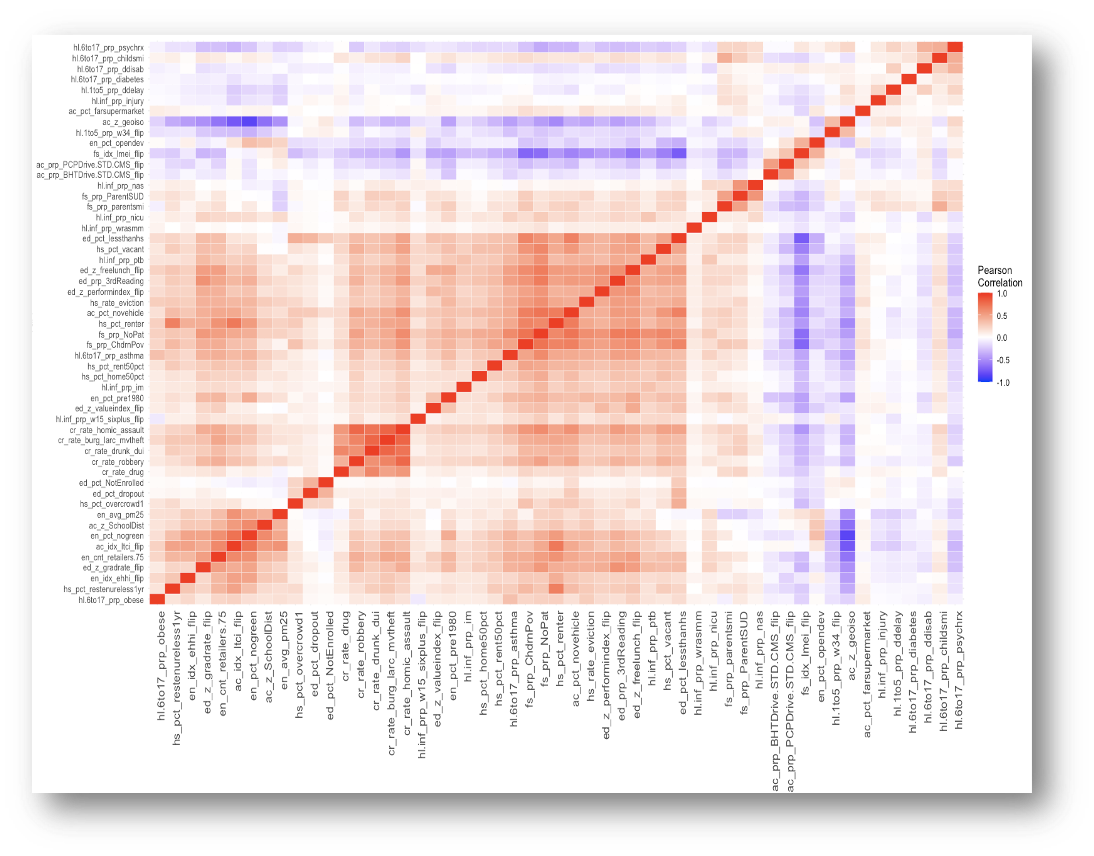

Constituent Measures

Constituent Measures

Figure S1. Heatmap with bivariate correlation values for the Ohio Children’s Opportunity Index (OCOI) constituent measures

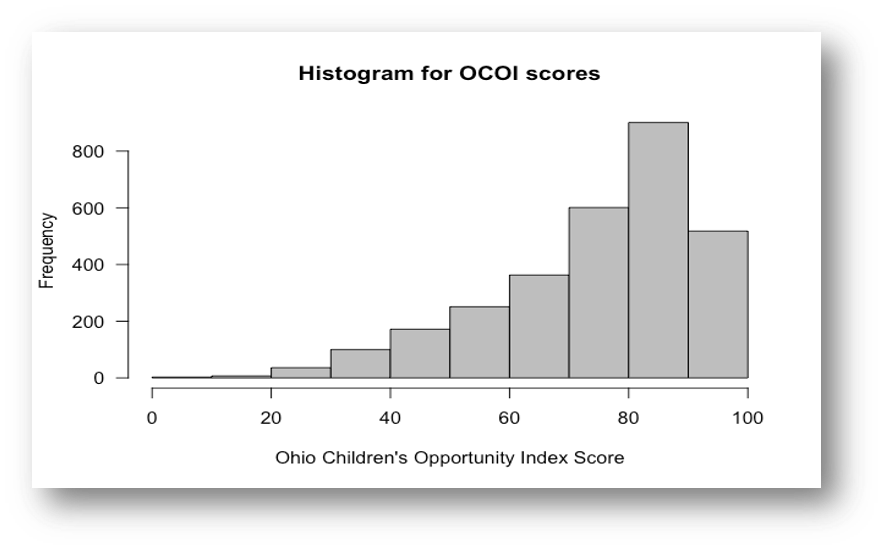

Figure S2. Distribution of Ohio Children’s Opportunity Index (OCOI) scores
